# Postnatal maternal depressive symptoms and behavioural outcomes in term- and preterm-born toddlers

**DOI:** 10.1101/2021.09.21.21263881

**Authors:** I Kleine, G Vamvakas, A Lautarescu, S Falconer, A Chew, SJ Counsell, A Pickles, AD Edwards, C Nosarti

## Abstract

**Objectives:** To examine the association between maternal depressive symptoms in the immediate postnatal period and offspring’s mental health in a large cohort of term- and preterm-born toddlers.

**Design and Participants:** Data were drawn from the Developing Human Connectome Project. Maternal postnatal depressive symptoms were assessed at term, and children’s outcomes were evaluated at a median corrected age of 18.4 months (range 17.3 – 24.3).

**Exposure and outcomes:** Preterm birth was defined as <37 weeks completed gestation. Maternal depressive symptoms were assessed with the Edinburgh Postnatal Depression Scale (EPDS). Toddlers’ outcome measures were parent-rated Child Behaviour Checklist 1^1/2^-5 Total (CBCL) and Quantitative Checklist for Autism in Toddlers (Q-CHAT) scores. Toddlers’ cognition was assessed with the Bayley Scales of Infant and Toddler Development – Third Edition (Bayley-III).

**Results:** Higher maternal EPDS scores were associated with toddlers’ higher CBCL (B=0.93, 95% CI 0.43-1.44, p<0.001, f^2^=0.05) and Q-CHAT scores (B=0.27, 95% CI 0.03-0.52, p=.031, f^2^=0.01). Higher maternal EPDS scores were not associated with toddlers’ cognitive outcomes. Maternal EPDS, toddlers’ CBCL and Q-CHAT scores did not differ between preterm (n=97; 19.1% of the total sample) and term participants.

**Conclusions:** Our findings indicate that children whose mothers had increased depressive symptoms in the early postnatal period, including subclinical symptoms, exhibit more maternally-reported behavioural problems in toddlerhood. These associations were independent of gestational age. Further research is needed to confirm the clinical significance of these findings.

**Strengths and limitations of this study:**

- Prospective study with a large sample, using multiple imputation to reduce non-response bias.
- Maternal depressive symptoms assessed as a continuous variable, providing more nuanced information about the significance of subclinical symptoms.
- Maternal depressive symptoms assessed earlier than in previous studies, enabling recognition of early screening opportunities for families.
- Potential shared method variance bias through parent-completed child behavioural assessments.
- Unknown paternal and parental factors, such as comorbid psychiatric conditions, that may confound our findings.

## Introduction

Postnatal depression affects approximately 12% of mothers worldwide.^1^ In contrast to ‘baby blues’, which is a state of emotional lability that affects between 30-80% of women in the first few days after birth and typically resolves spontaneously within two weeks,^2^ postnatal depression is more severe and starts in the first few months post-partum^1^. Stressful life events have been linked to a heightened risk of developing postnatal depression,^3^ with rates as high as 40% in women who give birth before term completion (i.e., preterm, < 37 gestational weeks),^4^ likely due to heightened stress associated with perinatal complications.^5^

Women with postnatal depression tend to be less responsive to their baby’s needs and to display less affection.^6^ Therefore, in the short-term postpartum depression may affect mother-infant interactions^7^ and in the long-term it may lead to alterations in brain development,^8^ emotional difficulties,^9^ less secure attachment, cognitive and behavioural problems in childhood, and a possible increased risk of autism spectrum disorder (ASD).^10,11^ Large cohort studies, such as the Avon Longitudinal Study of Parents and Children (ALSPAC), have shown that these associations are even evident when maternal depression is measured on a continuum of symptoms rather than a dichotomous diagnosis,^12–14^ supporting the notion that elevated sub-diagnostic psychiatric symptoms can also negatively impact on children’s development.^15^

Studies investigating the underlying causes that may link maternal postnatal depression to child outcomes have implicated several biological and environmental variables. For instance, genetic and epigenetic factors have been shown to both mediate and mitigate the intergenerational transmission of psychiatric disorders,^16^ while lower quality parenting, interparental conflict, and socioeconomic deprivation have been shown to exacerbate children’s developmental risk.^11^ In addition, preterm birth has been associated with alterations in early brain development,^17^ as well as neurological, behavioural and cognitive problems in childhood and beyond.^18,19^ Therefore, it is complex to disentangle the possible effects of postnatal maternal mental health and those of perinatal clinical factors on specific outcomes in preterm children, as these may involve both maternal psychosocial and biological variables and child preterm-related neurodevelopmental morbidity. Furthermore, a question that remains unanswered is whether perinatal clinical risk accentuates the association between maternal postnatal depressive symptoms and child outcome. Previous research has proposed a diathesis-stress model, whereby preterm birth is regarded as a vulnerability factor that makes preterm infants more prone to suboptimal environmental influences compared to term infants.^20,21^ On the other hand, the differential susceptibility model frames preterm birth as a plasticity factor that makes infants more likely to have both poorer outcomes in negative environments, as well as better outcomes in supportive environments.^21,22^

Given that mothers of preterm children experience elevated levels of distress,^23^ are at high risk of developing postnatal depression,^4^ and that preterm children themselves are vulnerable to psychiatric sequelae,^24^ we aimed to investigate the association between very early symptoms of maternal postnatal depression and child behavioural and emotional outcomes, as well as whether this association was influenced by gestational age. We hypothesise that early postnatal maternal depressive symptoms would be more elevated in mothers of preterm compared to term infants and that these would impact preterm children’s behavioural and emotional outcomes to a greater degree than their term counterparts.

## Methods

### Sample

Participants were enrolled in the Developing Human Connectome Project (DHCP, http://www.developingconnectome.org/). Toddlers were invited to the Centre for the Developing Brain, St Thomas’ Hospital, London, for neurodevelopmental assessment between 17 and 24 months post-expected delivery date. Inclusion criteria for our follow-up study were: mother and baby attendance for magnetic resonance imaging (MRI) at term corrected age; completed toddler neurodevelopmental assessment. 509 toddlers met these inclusion criteria by the date of closure for this analysis (26/02/2020). This study was approved by the UK National Research Ethics Authority (14/LO/1169) and conducted in accordance with the World Medical Association’s Code of Ethics (Declaration of Helsinki). Written informed consent was given by children’s carer(s) at recruitment into the study.

### Maternal variables

Maternal age, parity, Body Mass Index (BMI) and postcode were collected at enrolment into the DHCP study. Parity was coded as 0, 1, 2, or ≥3 previous children. Index of Multiple Deprivation (IMD) rank was computed from the current maternal postcode using the 2019 IMD classification,^25^ and provided a proxy for family socioeconomic status. Lower IMD rank corresponds to greater social deprivation.

*Maternal depressive symptoms* were measured using the Edinburgh Postnatal Depression Scale (EPDS)^26^ at term corrected age. The EPDS is a 10-item screening questionnaire completed by mothers, with higher scores reflecting a higher likelihood of depressive disorders. A score of 13 can be used as a cut-off indicating high-level symptoms, although a cut-off of 11 maximises the sensitivity and specificity of the screening tool for depression.^27^

### Child variables

Infant *clinical characteristics* included: sex, gestational age at birth, birth weight, and pregnancy size (singleton/twin/triplet).

*Behavioural outcomes* were assessed using the Child Behaviour Checklist/1^1/2^-5 (CBCL), a parent-completed 100-item questionnaire, in which the parent rates the child’s behaviour over the preceding two months using a 3-point Likert scale (“not true”, “somewhat or sometimes true”, and “very true or often true”). Responses are categorised into syndrome profiles, and these are subsequently grouped into internalising (emotional reactivity, anxiety/depression, somatic complaints, and withdrawal), externalising (attention problems, aggressive behaviour) and total (internalising, externalising, sleep and other) problem scales. Higher scores indicate increased emotional and behavioural problems. Total scores are classified into a normal range (≤92^rd^ centile, T ≤64), borderline range (93^rd^-97^th^ centile, T 65-69), and clinical range (≥98^th^ centile, T ≥70). The CBCL is known to have high reliability and validity for measuring children’s emotional and behavioural problems.^28^

*ASD traits* were assessed using the Quantitative Checklist for Autism in Toddlers (Q-CHAT). The Q-CHAT is a parent-completed 25-item questionnaire, in which the child’s behaviour is scored on a 5-point (0-4) frequency scale. Higher total scores correspond to a higher frequency of autistic traits. The Q-CHAT shows good test-retest reliability, face validity and specificity, yet poor positive predictive value.^29,30^

*Cognitive assessment* was performed using the Bayley Scales of Infant and Toddler Development – Third Edition (Bayley-III). The Bayley-III provides scores for a child’s overall cognitive, language and motor development. The cognitive standardised composite score was used in this study; scores between 70-85 indicate mild cognitive impairment, and scores lower than 70 indicate moderate-severe impairment^31^. Reliability and validity of the Bayley-III have been shown to be robust.^32^

Assessments were carried out by staff experienced in the neurocognitive assessments of toddlers.

### Analysis

Descriptive statistics and one-way ANOVA tests were performed in IBM SPSS Statistics for Windows v.25. All other analyses were carried out in Stata v.16.

Multiple imputation (MI) was carried out to account for missing data in CBCL (11/509, 2.24%), Q-CHAT (9/509, 1.8%), maternal EPDS (73/509, 14.3%), maternal BMI (27/509, 5.3%) and IMD rank (3/509, 0.6%). Variables were imputed simultaneously using the ‘mi impute chained’ procedure that performs imputation by chained equations. The imputation models included all variables that appear in the corresponding analysis models and also had the same structural form as the analysis models. They additionally included all variables correlating with the incomplete variables, as well as all predictors of the probability of a value being missing.^33^ Maternal depression and CBCL were imputed using Poisson regression; Q-CHAT, maternal BMI, and the IMD rank were imputed using linear regression. 40 MI datasets were created. To assess the stability of our MI parameters, we extracted the Monte Carlo error of each parameter estimate and examined whether the error for the coefficient was less than 10% of the parameter’s standard error estimate. MI estimates were used for the primary analyses and compared to the estimates from complete-case (CC, individuals who had no missing data pre-imputation) analyses. Normal probability plots of residuals from the CC analyses were examined.

The analysis models used multiple linear regression with standard errors that allowed for intragroup correlation and were fitted using the ‘mi estimate’ procedure, which estimates effects after application of Rubin’s rules.^34^ For continuous variables, Cohen’s f-squared effect sizes were calculated using 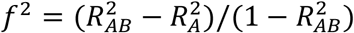, where 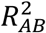 is the R-squared value from a regression model that includes the variable of interest as well as all the covariates used in the model, and 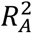 is the R-squared value from the regression model that includes only the covariates.^35,36^ For binary variables, Cohen’s f-squared effect sizes were produced after estimating first the Cohen’s d using the formula: 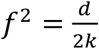, where k is the number of groups. As a measure of dispersion, Cohen’s d used the average root mean-square error over the MI datasets. Adjusted R-squared values after MI were extracted after estimating the model with the user-written ‘mibeta’ command with the ‘fisherz’ option,^37^ which calculates R-squared measures for linear regression with MI data. The significance of the joint effect of the categorical variable parity was assessed using ‘mi test’ which performs Wald tests of composite linear hypotheses.

*Primary outcome measures* were children’s total CBCL raw score and Q-CHAT score. S*econdary outcome measures* were CBCL internalising and externalising scores. The effect of maternal EPDS score was adjusted for IMD rank, maternal age, maternal BMI, maternal parity, pregnancy size, and the following child’s variables: gestational age, birth weight, Bayley-III cognitive composite score, and corrected age at assessment. The interaction between gestational age and maternal depression was explored using a complete case analysis in both CBCL and Q-CHAT models. EPDS, CBCL and Q-CHAT scores were compared between term and preterm infants using the complete case dataset.

In order to investigate the specificity of the association between maternal EPDS scores and child’s behavioural outcomes (versus cognitive outcomes) we repeated the analyses using the Bayley-III cognitive composite score as primary outcome, with the following confounders: IMD rank, maternal age, maternal BMI, maternal parity, pregnancy size, and the following child’s variables: gestational age, birth weight, corrected age at assessment, and Q-CHAT score. CBCL score was not included in the model predicting cognitive outcome, because cognition was not a significant predictor of CBCL (see Results).

As all mothers had their EPDS score measured near term-corrected age, we further investigated the association between time elapsing between baby’s birth and mother’s EPDS assessment and EPDS score, in order to avoid erroneously identifying ‘baby blues’ in mothers of term-born infants versus postnatal depression in mothers of preterm infants. This post-hoc analysis was performed using Poisson regression.

## Results

509 toddlers were followed up at a median corrected age of 18.4 months (range 17.3 – 24.3 months). 51 (10.0%) of these were twins, and 3 (0.59%) were triplets. 21/509 (4.13%) of mothers scored above a clinical cut-off (≥13) on the EPDS.(26) Demographic data are shown in Table 1. 400 (78.6%) children had complete data. Missing data were imputed and thus all 509 subjects were included in the primary and secondary analyses. One participant was excluded from the cognition analysis after examining the quintiles of the residuals against the theoretical quintiles of a normal distribution. The mean CBCL T score was 46.9 (SD 9.5) (Table 1); 484 (95.1%) of participants had a CBCL score in the normal range, 14 (2.8%) were borderline, and no participants scored in the clinical range. The mean Q-CHAT score was 30.5 (SD 9.3) (Table 1).

**Table 1:** Socio-demographic, maternal and clinical characteristics (n=509)

| **Variable** | **Number (%)\*** |
| --- | --- |
| Sex: Male | 274 (53.8) |
| Index of Multiple Deprivation (IMD) quintiles |  |
| 1 (least deprived)<sup>a</sup> | 65 (12.8) |
| 2 | 87 (17.2) |
| 3 | 108 (21.3) |
| 4 | 173 (34.2) |
| 5 (most deprived) | 73 (14.4) |
| Gestational age at birth (weeks), median [range] | 39.7 [20 – 43] |
| Gestational category |  |
| Preterm (<37 weeks) | 97 (19.1) |
| Term (≥37 weeks) | 412 (80.9) |
| Birthweight (g), median [range] | 3290 [450 – 4750] |
| Multiple pregnancy | 54 (10.6) |
| Maternal parity |  |
| 0 | 332 (65.2) |
| 1 | 124 (24.4) |
| 2 | 32 (6.3) |
| 3+ | 21 (4.2) |
| Maternal BMI (kg/m<sup>2</sup>), median [range] | 23.2 [15.3 – 43.6] |
| Maternal age at infant's birth (years), mean (SD) | 34.2 (4.8) |
| Bayley III cognitive composite score, mean (SD) | 100 (11.4) |
| CBCL total T score, mean (SD) | 46.9 (9.5) |
| Q-CHAT total score, mean (SD) | 30.5 (9.3) |
| EPDS score, median [range] | 4 [0 – 28] |

<sup>a</sup> Quintile 1 corresponds to the highest, least deprived, IMD rankings.
\* unless otherwise specified

Predictors of children’s CBCL and Q-CHAT scores after multiple imputation are shown in Table 2. Higher maternal EPDS score was associated with children’s higher CBCL Total score (B=0.93, 95% CI 0.43-1.44, p<0.001, f^2^=0.05) and Q-CHAT score (B=0.27, 95% CI 0.03-0.52, p=.031, f^2^=0.01) (Table 2). These associations are presented graphically in Figure 1 and Figure 2, respectively. Higher maternal EPDS score was associated with both internalising (B=0.22, 95% CI 0.08-0.36, p<0.01, f^2^=0.03) and externalising (B=0.40, 95% CI 0.20-0.61, p<0.001, f^2^=0.05) symptoms in children (Online Resource 1 and 2, respectively). Comparison of the imputed model analyses to the complete-case analyses showed that results were consistent for the CBCL model (Online Resource 3). Comparison for the Q-CHAT model showed that maternal EPDS was a significant predictor in the imputed model, but not in the complete-case analysis (Online Resource 3).

**Table 2:** CBCL and Q-CHAT model predictors using multiple imputation without interaction. (Cf. Online Resource 3 for complete case analysis)

|  | CBCL |  |  | Q-CHAT |  |  |
| --- | --- | --- | --- | --- | --- | --- |
|  | B [95%CI] | p | f<sup>2</sup> | B [95%CI] | p | f<sup>2</sup> |
| **Maternal EPDS** | 0.93 [0.43, 1.44] | <.001 *** | 0.05 | 0.27 [0.03, 0.52] | .031 * | 0.01 |
| **Maternal BMI** | -0.09 [-0.44, 0.26] | .621 | - | 0.06 [-0.13, 0.24] | .538 | - |
| **Multiple pregnancy** | 3.15 [-3.07, 9.37] | .320 | - | 1.33 [-2.62, 5.28] | .509 | - |
| **Parity** |  |  |  |  |  |  |
| **1** | -2.52 [-5.96, 0.93] | .151 | - | -2.14 [-4.02, -0.27] | .025 <sup>a</sup> | - |
| **2** | -3.23 [-9.16, 2.70] | .285 | - | 0.88 [-1.99, 3.75] | .548 | - |
| **3+** | -1.37 [-8.36, 5.61] | .699 | - | -0.49 [-4.57, 3.60] | .815 | - |
| **IMD rank** | -1.48 [-3.33, 0.37] | .117 | - | -1.50 [-2.60, -0.40] | .008 ** | 0.02 |
| **Gestational age at birth (weeks)** | 0.10 [-0.65, 0.85] | .786 | - | 0.26 [-0.17, 0.70] | .233 | - |
| **Birthweight (kg)** | 0.56 [-2.65, 3.78] | .731 | - | -1.24 [-2.93, 0.46] | .151 | - |
| **Sex:female** | -4.14 [-6.96, -1.31] | .004 ** | 0.06 | -1.95 [-3.42, -0.48] | .009 ** | 0.05 |
| **Corrected age at assessment (months)** | -0.90 [-2.17, 0.37] | .166 | - | -0.16 [-0.91, 0.59] | .677 | - |
| **Cognition** | -0.05 [-0.20, 0.09] | .467 | - | -0.27 [-0.35, -0.20] | <.001 *** | 0.12 |

p<0.05 \*; p<0.01 \*\*; p<0.001 \*\*\*
CBCL model adjusted R<sup>2</sup> = 0.0676. Q-CHAT model adjusted R<sup>2</sup> = 0.193.
B = unstandardised coefficient.
CBCL = Child Behaviour Checklist score at 18 months. Q-CHAT = Quantitative Checklist for Autism in Toddlers score at 18 months. Maternal EPDS = maternal Edinburgh Postnatal Depression Scale score at term-corrected age. Multiple pregnancy = dummy variable of twin/triplet pregnancy. Parity = dummy variable, one/two/three+ previous child(ren). Corrected age at assessment (months) = age at behavioural assessment, corrected for gestational age. Cognition = infant Bayley III score at 18 months.
<sup>a</sup>Wald test of whole parity variable in Q-CHAT model: F(3, 476.9) = 1.88, p = .133
Effect size (Cohen's f<sup>2</sup>, calculated from squared part correlations for predictors significant to 0.05): 0.02 = small, 0.15 = medium and 0.35 = large.[25]
- indicates data not given, as predictor not significant to 0.05.

**Fig. 1.**
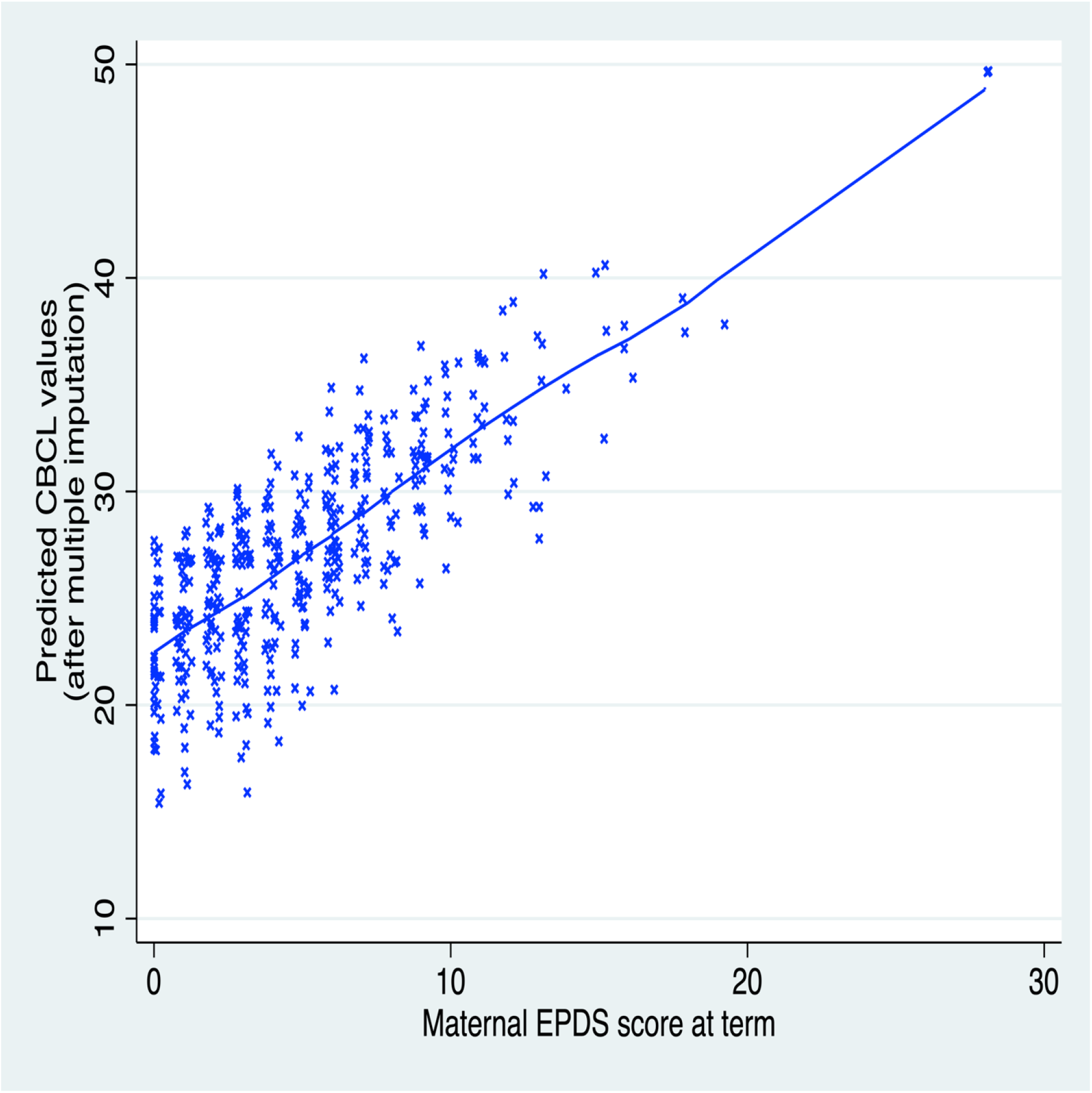
Children’s predicted CBCL scores at 18 months are positively correlated to the maternal EPDS score at term corrected age

**Fig. 2.**
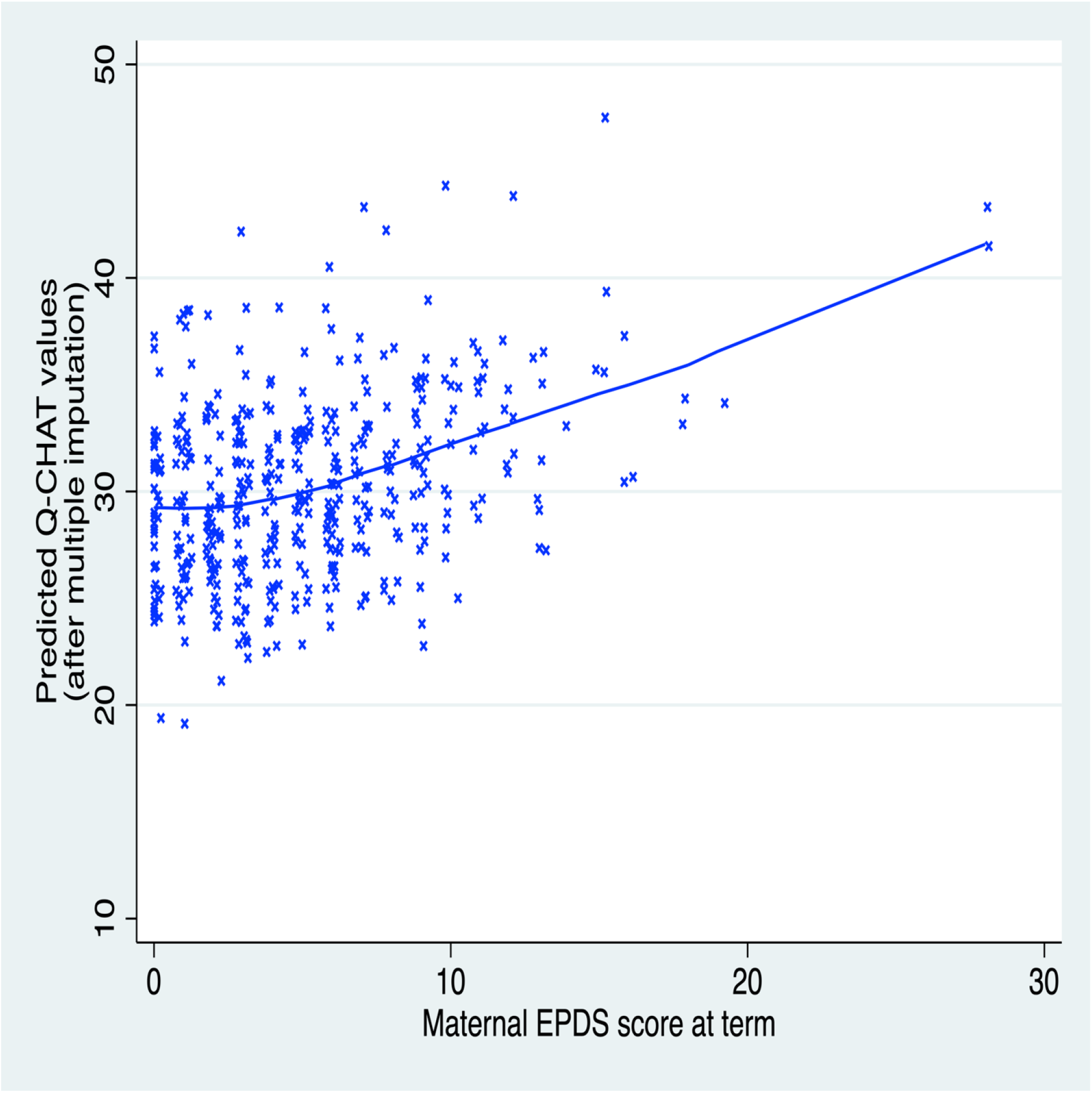
Children’s predicted Q-CHAT scores at 18 months are positively correlated to the maternal EPDS score at term corrected age

Maternal EPDS scores did not differ between preterm and term groups in the complete dataset (t(434)=0.11, p=0.92). CBCL scores (t(496)=0.95, p=0.34) and Q-CHAT scores (t(122.6)=0.50, p=0.62) did not differ between preterm and term groups in the complete dataset. Maternal EPDS score did not disproportionately affect preterm children with respect to CBCL or Q-CHAT scores (Table 3).

**Table 3:** CBCL and Q-CHAT model predictors using complete case analysis with interaction of ‘EPDS x term’.

|  | CBCL |  | Q-CHAT |  |
| --- | --- | --- | --- | --- |
|  | B [95%CI] | p | B [95%CI] | p |
| **Maternal EPDS** | 0.89 [-0.24, 2.02] | .121 | 0.24 [-0.28, 0.75] | .365 |
| **Maternal BMI** | -0.01 [-0.39, 0.37] | .955 | 0.00 [-0.16, 0.17] | .982 |
| **Multiple pregnancy** | 1.76 [-6.65, 10.17] | .681 | 0.97 [-2.07, 4.01] | .532 |
| **Parity** |  |  |  |  |
| **1** | -2.75 [-6.49, 0.99] | .149 | -1.42 [-3.30, 0.46] | .139 |
| **2** | -3.49 [-10.36, 3.37] | .317 | 0.16 [-2.84, 3.16] | .917 |
| **3+** | -1.17 [-9.69, 7.35] | .788 | -1.13 [-4.24, 1.98] | .476 |
| **IMD rank** | -1.41 [-3.54, 0.73] | .195 | -1.68 [-2.64, -0.72] | .001 ** |
| **Gestation: term** | 1.25 [-8.34, 10.85] | .797 | 2.64 [-1.74, 7.02] | .236 |
| **Birthweight (kg)** | -1.01 [-4.08, 2.05] | .516 | -2.25 [-3.73, -0.78] | .003 ** |
| **Sex: female** | -4.64 [-7.83, -1.44] | .005 ** | -2.22 [-3.72, -0.71] | .004 ** |
| **Corrected age at assessment (months)** | -0.83 [-2.27, 0.62] | .261 | -0.39 [-1.18, 0.04] | .335 |
| **Cognition** | -0.03 [-0.20, 0.14] | .720 | -0.22 [-0.29, -0.15] | <.001 *** |
| **EPDS x gestation:term** | -0.01 [-1.30, 1.28] | .991 | -0.02 [-0.60, 0.56] | .950 |

p<0.05 \*; p<0.01 \*\*; p<0.001 \*\*\*
CBCL model adjusted $R^2 = 0.0865$ . Q-CHAT model adjusted $R^2 = 0.215$ .
B = unstandardised coefficient.
CBCL = Child Behaviour Checklist score at 18 months. Q-CHAT = Quantitative Checklist for Autism in Toddlers score at 18 months. Maternal EPDS = maternal Edinburgh Postnatal Depression Scale score at term-corrected age. Multiple pregnancy = dummy variable of twin/triplet pregnancy. Parity = dummy variable, one/two/three+ previous child(ren). Gestation: term = dummy variable, term ( $\geq 37$ weeks) vs preterm ( $< 37$ weeks) gestation at birth. Corrected age at assessment (months) = age at behavioural assessment, corrected for gestational age. Cognition = infant Bayley III score at 18 months. EPDS x gestation:term = interaction term between maternal EPDS score and term gestation at birth.

Mothers who gave birth prematurely (<37 weeks gestation) had their EPDS score assessed on average 7.7 weeks later post-delivery than mothers who gave birth at term (preterm participants M=8.9 (SD 4.8), term participants M=1.2 (SD 1.3); t(99.4)=15.5, p<.001). The time-lag between birth and EPDS assessment did not predict maternal EPDS score, and there was no evidence of a significant interaction between gestation and birth-to-assessment time-lag (Online Resource 4 and 5, respectively).

Boys had higher CBCL and Q-CHAT scores than girls. Higher Q-CHAT scores were associated with lower IMD rank (i.e., greater socio-economic deprivation) and lower Bayley-III cognitive composite scores. Parity was not a significant predictor of outcome in any of the models (Table 2).

The mean Bayley III cognitive composite score in our sample was 100 (SD 11.4) (Table 1); this corresponds to the standardised test mean.^31^ 480 (94.3%) of participants had a normal cognitive score, 24 (4.7%) had mild impairment, and 5 (1%) had moderate-severe impairment. Predictors of children’s cognitive score are shown in Table 4. Maternal EPDS score at term was not associated with toddlers’ cognitive outcomes (B=-0.22, 95% CI -0.50-0.05, p=.108) (Table 4).

**Table 4:** Cognition model predictors using multiple imputation.

|  | **B [95%CI]** | **p** |
| --- | --- | --- |
| **Maternal EPDS** | -0.22 [-0.50, 0.05] | .108 |
| **Maternal BMI** | -0.32 [-0.52, -0.13] | .001 ** |
| **Multiple pregnancy** | 1.65 [-2.49, 5.79] | .433 |
| **Parity** |  |  |
| **1** | -0.46 [-2.67, 1.76] | .686 |
| **2** | -3.47 [-6.69, -0.25] | .035 <sup>a</sup> |
| **3+** | -4.57 [-9.53, 0.40] | .072 |
| **IMD rank** | 1.43 [0.37, 2.50] | .009 ** |
| **Gestational age at birth (weeks)** | 0.45 [-0.07, 0.96] | .091 |
| **Birthweight (kg)** | 0.81 [-1.24, 2.87] | .436 |
| **Sex: female** | 1.99 [0.24, 3.74] | .026 * |
| **Corrected age at assessment (months)** | -0.75 [-1.59, 0.08] | .075 |
| **Q-CHAT score** | -0.39 [-0.50, -0.28] | <.001 *** |

p<0.05 \*; p<0.01 \*\*; p<0.001 \*\*\*
Adjusted R<sup>2</sup> = 0.231
B = unstandardised coefficient.
Maternal EPDS = maternal Edinburgh Postnatal Depression Scale score at term-corrected age.
Multiple pregnancy = dummy variable of twin/triplet pregnancy. Parity = dummy variable, one/two/three+ previous child(ren). Corrected age at assessment (months) = age at behavioural assessment, corrected for gestational age. Cognition = infant Bayley III score at 18 months. Q-CHAT score = infant's Q-CHAT score at 18 month assessment.
<sup>a</sup> Wald test of whole parity variable in cognition model: F(3, 482.9)=2.41, p=0.067

## Discussion

### Principal findings

Contrary to our predictions, mothers of preterm infants did not display more depressive symptoms compared to mothers of term infants. Moreover, gestational age not influence the association between maternal depressive symptoms and infants’ behavioural outcomes in toddlerhood. These results suggest that preterm birth may not be a vulnerability or plasticity factor with respect to the effect of maternal postnatal depression on infants’ behavioural development in the first 18 months of life. However, our results do suggest that more maternal self-reported depressive symptoms shortly after birth are associated with greater toddlers’ behavioural problems and ASD traits, but not with cognitive outcomes. Given that fewer than 5% of the mothers in our cohort had EPDS scores above a clinical threshold,^26^ our findings indicate that even subclinical depressive symptoms adversely impact children’s behavioural outcomes. In addition, our cohort was typically developing with few CBCL scores reaching a concerning threshold; our results could be interpreted within the conceptual framework of mental illness lying on a continuum with typical behavioural traits.^38^

### Comparison to prior literature

The finding that more preterm infants were not disproportionately affected by maternal depressive symptoms supports Hadfield et al.’s findings that maternal distress at 9 months did not differentially impact very preterm (<34 weeks) or late preterm (34-36^+6^ weeks) infants with respect to socioemotional outcomes, although paternal distress did have an impact on very preterm infants’ outcomes.^39^ However, our results differ from Gueron-Sela et al.’s finding that very preterm (28-33 weeks) 12 month old infants’ social outcomes were more influenced by maternal emotional distress at 6 months than term infants’ outcomes.^22^ The inconsistent findings may be due to methodological differences: for instance, our infant assessment being conducted at 18 months corrected age when social competency is more developed, our assessment of maternal depressive symptoms being in the very early postnatal period, or our use of a screening measure, the Q-CHAT, as a measure of ASD traits. Importantly, the lack of support for a diathesis-stress or differential susceptibility model of maternal mental state on younger preterm infants in our study must be viewed in the context of our results also showing no difference in CBCL and Q-CHAT scores between term and preterm infants. This is in contrast to the existing literature that preterm infants are more likely to develop behavioural problems, such as ADHD, in childhood and adolescence.^19,24^ It is possible that the phenotypes of neurodevelopmental and neuropsychiatric disorders assessed with the chosen behavioural measures may not be sufficiently expressed at 18 months corrected age.^40^ In addition, as briefly discussed above, much of the existing literature emphasises the risk of extreme (<28 weeks) or very preterm (28-33 weeks) birth on later mental health outcomes,^19,24^ whereas only 3.5% and 5.5% of our participants fell within the extreme and very preterm birth group, respectively, and we thus may not have the power to show any subtle effects.

Our results with respect to internalising and externalising behavioural outcomes are in line with previous studies, including large population cohort studies, that show an association between postnatal maternal depression and young children’s emotional and behavioural problems.^11^ The only previous study investigating this association in infants at 18 months found maternal depression to be associated with internalising and dysregulated behaviour, but not externalising symptoms.^41^ This difference between our and Conroy et al.’s findings may have arisen from their exclusion of infants born <36 weeks and their use of a clinical diagnosis of depression for mothers, rather than the dimensional approach we employed. Interestingly, our finding that even subclinical depressive symptoms may adversely impact children’s behavioural outcomes is in line with recent data showing that low-as well as high-level depressive symptoms are associated with internalising and externalising symptoms in children aged 3 years.^42^

The results showing an association between maternal postnatal depressive symptoms and childhood ASD are less robust and need to be interpreted with caution. Although some prior studies have reported an association between antenatal maternal depression and offspring’s ASD,^10,43^ and postnatal depression has been suggested as a potential focus of cross-domain studies of ASD,^44^ there is no clear aetiological role of maternal postnatal depression in the development of ASD *per se*. Also, given that mothers with ASD are more likely to suffer from perinatal depression than mothers without ASD,^45^ and ASD is highly heritable,^46^ maternal depression may be a confounder in our observed results.

### Strengths & limitations of the study

The strengths of this study lie primarily in its large sample and prospective data collection. Moreover, the use of multiple imputation methodology has facilitated retention of a complete dataset, thus minimising non-response bias and increasing parameter precision. A strength in comparison to prior population cohort studies is that we assessed very early maternal depressive symptoms. Given the complex interplay of biological and environmental factors in the aetiology of mental health disorders, the inclusion of a substantive proportion of preterm infants in our cohort also offers an important insight into the role of preterm birth in influencing mental health outcomes; moreover, our results represent the full gestational spectrum, rather than discrete gestational categories. In addition, using maternal depression as a continuous, rather than dichotomous, variable allows a more nuanced understanding of the role maternal postnatal depressive symptoms may play in influencing children’s outcomes.

There are five main limitations to this study. Firstly, differences in birth-to-EPDS-assessment time-lags are a potential confounder, given the time-sensitive nature of early-onset temporary baby blues and later-onset pathological postnatal depression. Mothers of infants born at term were assessed early post-delivery, within the period one would anticipate baby blues to present, whereas mothers of preterm participants were on average assessed later, when postnatal depression predominates.^1,2^ Although our post-hoc analyses showed no association between the time elapsed from birth to EPDS assessment and maternal EPDS score, providing reassurance that our assessments of mothers of term-born infants were not inflated by the common, temporary symptoms of baby blues, it is however possible that we did not capture the full extent of later-onset depressive symptoms in mothers of term-born infants. This may explain why maternal EPDS scores did not differ between preterm and term groups in our complete dataset analysis, contrary to the current literature.^23^ Secondly, a number of important confounders that are likely to affect children’s behavioural outcomes were not assessed in this study, including genetic risk for psychiatric disorders,^47^ parental psychiatric co-morbidities,^41^ chronicity of postnatal depressive symptoms,^42^ antenatal maternal depression, paternal depression and subsequent parent-infant attachment, and inter-parental conflict.^11^ In this study we did not systematically collect maternal psychiatric history and our focus was on symptoms rather than a diagnosis of depression. Thus, we are unable to conclude whether our observed associations between early postnatal maternal depressive symptoms and children’s mental health outcomes are moderated or mediated by other parental factors. Thirdly, whilst our study included a substantive proportion of preterm infants (97/509, 19%), the sample was not random, as preterm children were selectively recruited for the DHCP; indeed, preterm infants are over-represented in our sample when compared to the UK population incidence (7.3%),^48^ which may limit the study’s generalisability to the general population. Fourthly, the effect sizes of the association between maternal EPDS score and behavioural problems and ASD traits, respectively, were small; this raises questions regarding the clinical significance of our findings and potentially explains some of the inconsistency between this and previous studies. Even within our analyses, the association between maternal depressive symptoms and ASD traits was not observed in our complete case analysis, thus calling into question the validity of this result. It is also important to highlight the continuum of ASD traits that are conceptualised by the Q-CHAT,^29^ as well as its poor positive predictive value;^30^ the presence of traits does not imply a diagnosis of ASD, and this distinction may also explain the contrast to previous studies. Fifthly, the outcome measures used in this study were parent-completed questionnaires and it is possible that reporting bias with shared method variance may have skewed our results, as maternal depression has been shown to influence reporting of ASD traits,^49^ including the Q-CHAT,^50^ and CBCL scores.^51^

### Implications of our findings

Of greatest importance to clinicians and policymakers is our finding that even *subclinical* maternal depressive symptoms are associated with behavioural outcomes of offspring. This has significant implications for the risk-stratification of women and their babies in the postnatal period, during which contact with medical professionals is already established. Identifying high risk families and providing appropriate supportive measures at the early postnatal stage may help to prevent future psychiatric morbidity.

### Future research

Further follow-up of large cohorts of preterm and term infants, to an age when behavioural phenotypes may become more pronounced, is needed to investigate whether the long-term developmental trajectories of term and ex-preterm infants are differentially susceptible to changes of postnatal maternal mental health. Such follow-up should use independent, objective assessments of child behavioural outcomes. Further study is also needed to elucidate the role of maternal depression in the aetiology of ASD, controlling for both diagnostic and sub-clinical maternal ASD symptomatology. Finally, it is crucial for future research to elucidate the interplay of biochemical and neurodevelopmental changes that may mediate and confound the translation of environmental exposures into distal behavioural phenotypes.

### Conclusion

This prospective longitudinal cohort study found no evidence to support the concept of preterm birth as a vulnerability or plasticity factor with respect to the effect of maternal depressive symptoms on behavioural development. However, we do show that early subclinical maternal postnatal depressive symptoms are associated with behavioural problems in children. This adds to the increasing body of literature indicating the role of subclinical and early postnatal depressive symptoms in the aetiology of childhood mental health disorders.

These findings are of great relevance to child and public health, and have potentially significant implications for developing strategies to facilitate effective screening and support for women and children, enabling all to reach their full potential.

## Data Availability

Research data will be available as part of Developing Human Connectome Project.

http://www.developingconnectome.org/

## Declarations

### Availability of data and material

Research data will be available as part of Developing Human Connectome Project (http://www.developingconnectome.org/).

### Code availability

Not applicable.

### Ethics approval

This study was approved by the UK National Research Ethics Authority (14/LO/1169) and conducted in accordance with the World Medical Association’s Code of Ethics (Declaration of Helsinki).

### Consent to participate

Written informed consent was given by children’s carer(s) at recruitment into the study.

### Consent for publication

Not applicable.

### Patient and Public Involvement statement

The current study was developed in consultation with the Weston Programme for Family Centered Research, which involves parents to define what research is valuable to them, and to allow them to lead it with support from the scientists in the Centre for the Developing Brain.

## Acknowledgements & contributions

We thank all DHCP investigators for their contribution to the study. We thank Dr Oliver Gale-Grant MRes (Centre for the Developing Brain, King’s College London; Department of Forensic & Neurodevelopmental Sciences, King’s College London) for providing the IMD rank data. We are very grateful to the families who generously took part in this research.

**Conceptualization:**SC, ADE, CN; **Methodology:** IK, GV, SC, AP, ADE, CN; **Investigation:** SF, AC; **Data curation:** IK, AL; **Formal analysis:** IK, GV, AP, CN; **Writing – original draft preparation:** IK; **Writing – Review & Editing:** GV, AL, SF, AC, SC, AP, ADE, CN; **Visualisation:** IK, GV; **Funding acquisition:** SC, ADE; **Supervision:** AP, ADE, CN.

