## Supplemental material for "Postnatal maternal depressive symptoms and behavioural outcomes in term- and preterm-born toddlers"

**Online Resource 1:** CBCL internalising symptom model predictors using multiple imputation

**Online Resource 2:** CBCL externalising symptom model predictors using multiple imputation

**Online Resource 3:** CBCL and Q-CHAT model predictors using complete case analysis without interaction.

**Online Resource 4:** EPDS score predictors

**Online Resource 5:** EPDS score predictors including interaction ‘term x time-lag’

**Online Resource 1:** CBCL internalising symptom model predictors using multiple imputation

| | B [95%CI] | p | $f^2$ |
|---|---|---|---|
| **Maternal EPDS** | 0.22 [0.08, 0.36] | .003 ** | 0.03 |
| **Maternal BMI** | -0.04 [-0.13, 0.06] | .436 | - |
| **Multiple pregnancy** | 0.58 [-1.10, 2.27] | .497 | - |
| **Parity** | | | |
| **1** | -0.37 [-1.41, 0.67] | .487 | - |
| **2** | -1.33 [-3.09, 0.42] | .136 | - |
| **3+** | 0.64 [-1.34, 2.62] | .524 | - |
| **IMD rank** | -0.41 [-0.91, 0.10] | .115 | - |
| **Gestational age at birth (weeks)** | 0.22 [-0.00, 0.44] | .053 | - |
| **Birthweight (kg)** | -0.97 [-1.90, -0.05] | .038 * | 0.005 |
| **Sex: female** | -0.51 [-1.35, 0.33] | .232 | - |
| **Corrected age at assessment (months)** | 0.02 [-0.41, 0.45] | .923 | - |
| **Cognition** | -0.05 [-0.10, -0.01] | .016 * | 0.01 |

$p<0.05$ *; $p<0.01$ **; $p<0.001$ ***

Adjusted $R^2 = 0.0566$.

B = unstandardised coefficient.

Outcome variable = Child Behaviour Checklist internalising sub-score at 18 months. Maternal EPDS = maternal Edinburgh Postnatal Depression Scale score at term-corrected age. Multiple pregnancy = dummy variable of twin/triplet pregnancy. Parity = dummy variable, one/two/three+ previous child(ren). Corrected age at assessment (months) = age at behavioural assessment, corrected for gestational age. Cognition = infant Bayley III score at 18 months.

Effect size (Cohen's $f^2$, calculated from squared part correlations for predictors significant to 0.05): 0.02 = small, 0.15 = medium and 0.35 = large.[1]

- indicates data not given, as predictor not significant to 0.05.

**Online Resource 2:** CBCL externalising symptom model predictors using multiple imputation

| | B [95%CI] | p | $f^2$ |
|---|---|---|---|
| **Maternal EPDS** | 0.40 [0.20, 0.61] | <.001 *** | 0.05 |
| **Maternal BMI** | 0.01 [-0.17, 0.15] | .933 | - |
| **Multiple pregnancy** | 2.51 [-0.29, 5.31] | .079 | - |
| **Parity** | | | |
| **1** | -1.06 [-2.53, 0.42] | .160 | - |
| **2** | -0.61 [-3.55, 2.33] | .682 | - |
| **3+** | -0.96 [-4.47, 2.56] | .593 | - |
| **IMD rank** | -0.24 [-1.11, 0.63] | .585 | - |
| **Gestational age at birth (weeks)** | -0.07 [-0.40, 0.27] | .701 | - |
| **Birthweight (kg)** | 1.03 [-0.38, 2.44] | .153 | - |
| **Sex: female** | -1.80 [-3.07, -0.53] | .006 ** | 0.06 |
| **Corrected age at assessment (months)** | -0.40 [-0.95, 0.16] | .161 | - |
| **Cognition** | 0.03 [-0.03, 0.10] | .322 | - |

p<0.05 *; p<0.01 **; p<0.001 ***
Adjusted $R^2$ = 0.0612.
B = unstandardised coefficient.
Outcome variable = Child Behaviour Checklist externalising sub-score at 18 months. Maternal EPDS = maternal Edinburgh Postnatal Depression Scale score at term-corrected age. Multiple pregnancy = dummy variable of twin/triplet pregnancy. Parity = dummy variable, one/two/three+ previous child(ren). Corrected age at assessment (months) = age at behavioural assessment, corrected for gestational age.
Cognition = infant Bayley III score at 18 months.
Effect size (Cohen's $f^2$, calculated from squared part correlations for predictors significant to 0.05): 0.02 = small, 0.15 = medium and 0.35 = large.[1]
- indicates data not given, as predictor not significant to 0.05.

**Online resource 3:** CBCL and Q-CHAT model predictors using complete case analysis without interaction.

| | CBCL | | Q-CHAT | |
|---|---|---|---|---|
| | **B [95%CI]** | **p** | **B [95%CI]** | **p** |
| **Maternal EPDS** | 0.88 [0.35, 1.41] | .001 ** | 0.21 [-0.02, 0.44] | .069 |
| **Maternal BMI** | -0.01 [-0.38, 0.37] | .963 | 0.01 [-0.16, 0.17] | .930 |
| **Multiple pregnancy** | 1.50 [-6.87, 9.87] | .724 | 0.43 [-2.50, 3.37] | .772 |
| **Parity** | | | | |
| **1** | -2.83 [-6.60, 0.94] | .141 | -1.54 [-3.41, 0.34] | .108 |
| **2** | -3.49 [-10.3, 3.35] | .316 | 0.13 [-2.85, 3.10] | .933 |
| **3+** | -1.38 [-10.0, 7.30] | .755 | -1.43 [-4.50, 1.64] | .360 |
| **IMD rank** | -1.44 [-3.56, 0.67] | .181 | -1.75 [-2.70, -0.79] | <.001 *** |
| **Gestational age at birth (weeks)** | 0.01 [-0.90, 0.91] | .987 | 0.10 [-0.33, 0.54] | .639 |
| **Birthweight (kg)** | -0.65 [-4.38, 3.08] | .733 | -1.81 [-3.63, 0.00] | .050 |
| **Sex: female** | -4.57 [-7.82, -1.31] | .006 ** | -2.12 [-3.60, -0.64] | .005 ** |
| **Corrected age at assessment (months)** | -0.84 [-2.26, 0.59] | .247 | -0.41 [-1.18, 0.35] | .290 |
| **Cognition** | -0.03 [-0.20, 0.13] | .689 | -0.23 [-0.30, -0.15] | <.001 *** |

p<0.05 *; p<0.01 **; p<0.001 ***
CBCL adjusted $R^2$ = 0.0862. Q-CHAT adjusted $R^2$ = 0.2103.
B = unstandardised coefficient.
CBCL = Child Behaviour Checklist externalising sub-score at 18 months. Q-CHAT = Quantitative Checklist for Autism in Toddlers score at 18 months. Maternal EPDS = maternal Edinburgh Postnatal Depression Scale score at term-corrected age. Multiple pregnancy = dummy variable of twin/triplet pregnancy. Parity = dummy variable, one/two/three+ previous child(ren). Gestation (weeks) = dummy variable: 34-36+6 weeks and ≥37 weeks gestation at birth. Corrected age at assessment (months) = age at behavioural assessment, corrected for gestational age. Cognition = infant Bayley III score at 18 months.

**Online Resource 4:** EPDS score predictors

| | IRR [95%CI] | p |
|---|---|---|
| **Time-lag (weeks)** | 1.01 [0.97, 1.05] | .647 |
| **Gestation:term** | 0.91 [0.64, 1.31] | .627 |
| **IMD rank** | 1.00 [1.00, 1.00] | .103 |
| **Multiple pregnancy** | 0.66 [0.46, 0.96] | .031 * |
| **Parity** | | |
| **1** | 0.79 [0.66, 0.95] | .011 * |
| **2** | 0.87 [0.60, 1.28] | .491 |
| **3+** | 0.84 [0.54, 1.31] | .445 |
| **Birthweight (kg)** | 0.98 [0.83, 1.17] | .847 |
| **Sex:female** | 1.13 [0.98, 1.31] | .098 |

p<0.05 *; p<0.01 **; p<0.001 ***

Pseudo $R^2$ = 0.0228

IRR = incidence rate ratio

Outcome variable = maternal Edinburgh Postnatal Depression Scale (EPDS) score at term-corrected age. Time-lag (weeks) = time in weeks between birth and EPDS assessment. Gestation:term = dummy variable, term (≥37 weeks) vs preterm (<37 weeks) gestation at birth. Multiple pregnancy = dummy variable of twin/triplet pregnancy. Parity = dummy variable, one/two/three+ previous child(ren).

**Online Resource 5:** EPDS score predictors including interaction 'term x time-lag'

| | IRR [95%CI] | p |
|---|---|---|
| **Time-lag (weeks)** | 1.00 [0.97, 1.04] | .823 |
| **Gestation: term** | 0.88 [0.58, 1.34] | .553 |
| **IMD rank** | 1.00 [1.00, 1.00] | .104 |
| **Multiple pregnancy** | 0.66 [0.46, 0.96] | .029 * |
| **Parity** | | |
| **1** | 0.79 [0.66, 0.95] | .010 * |
| **2** | 0.87 [0.60, 1.28] | .480 |
| **3+** | 0.85 [0.55, 1.31] | .458 |
| **Birthweight (kg)** | 0.97 [0.83, 1.15] | .756 |
| **Sex:female** | 1.13 [0.98, 1.30] | .100 |
| **Term x time-lag (weeks)** | 1.01 [0.94, 1.10] | .735 |

$p<0.05$ *; $p<0.01$ **; $p<0.001$ ***
Pseudo $R^2$ = 0.0230
IRR = incidence rate ratio
Outcome variable = maternal Edinburgh Postnatal Depression Scale (EPDS) score at term-corrected age. Time-lag (weeks) = time in weeks between birth and EPDS assessment. Gestation: term = dummy variable, term (≥37 weeks) vs preterm (<37 weeks) gestation at birth. Multiple pregnancy = dummy variable of twin/triplet pregnancy. Parity = dummy variable, one/two/three+ previous child(ren). Term x time-lag (weeks): interaction term between term gestation at birth and time-lag between birth and maternal EPDS assessment.
